# Thoracic Computed Tomographic Abnormalities in Patients with Spinal Tuberculosis: A Prospective Evaluation

**DOI:** 10.1101/2024.11.06.24316827

**Authors:** Anil Kumar Sahu, Neeraj Kumar, Ravindra Kumar Garg, Hardeep Singh Malhotra, Kiran Preet Malhotra, Imran Rizvi, Amita Jain, Anit Parihar, Rajesh Verma, Praveen Kumar Sharma, Ravi Uniyal, Shweta Pandey

**Author notes:** The first three authors have contributed equally. Address for correspondence Ravindra Kumar Garg, Department of Neurology, King George Medical University, Uttar Pradesh, Lucknow, India PIN-226003.

## Abstract

**Background:** Spinal tuberculosis is the most common osteoarticular tuberculosis. Many published isolated reports indicated that many patients with spinal tuberculosis have lung abnormalities. In this study, we aimed to prospectively evaluate the prevalence and spectrum of lung abnormalities.

**Methods:** We enrolled consecutive newly diagnosed patients with spinal tuberculosis. They were subjected to spinal magnetic resonance imaging and computed tomography (CT) of the chest. They were followed for 6 months. The outcome was assessed by using the modified Rankin scale.

**Results:** Out of 66 patients, 22 (33.3%) patients had CT thorax abnormalities. The most frequent finding was lung fibrosis (13 patients). Six patients had consolidation. Miliary tuberculosis was demonstrated in one patient. Cavitation, atelectasis, emphysema, and traction bronchiectasis were noted in one patient each. Nine patients had pleural effusion. Pleural thickening and empyema were noted in two patients each. Empyema was noted in 2 patients. Hydropneumothorax was seen in one patient. One patient had endobronchial tuberculosis. Hilar and Mediastinal lymphadenopathy was noted in 6 patients. At inclusion, 59 (89.4%) patients had modified Rankin scale scores≥3. After 6 months, 58 (87.9%) patients showed improvement and achieved a score <3. In 4 patients, spinal surgery was needed. The remaining three patient’s disability statuses remained unaltered. CT chest abnormalities were not associated with improvement in disability.

**Conclusion:** One-third of patients with spinal tuberculosis had tuberculous CT thorax abnormalities. The most frequent CT thorax finding was lung fibrosis. CT abnormalities did not affect the outcome.

## Introduction

Spinal tuberculosis is the most common form of musculoskeletal tuberculosis. It constitutes about 50% of skeletal tuberculosis cases. Held and colleagues in their review mentioned that approximately 10-25% of extrapulmonary tuberculosis is musculoskeletal tuberculosis.^1^ Between 2002 and 2011 in the USA, a total of 75,858 tuberculosis patients were recorded, of whom 2789 (3.7%) had spinal tuberculosis.^2^ Spinal tuberculosis is much more common in high tuberculosis burden countries, however, the precise incidence is not known.

In the pathogenesis of spinal tuberculosis, *Mycobacterium tuberculosis* lung infection is the first event. *Mycobacterium tuberculosis* spreads to the lungs via an airborne route, through droplet nuclei containing *Mycobacterium tuberculosis.* Spinal tuberculosis usually develops as a result of the hematogenous spread of *Mycobacterium tuberculosis* to vertebral structures. High vascularity of the vertebral marrow enhances the risk of *Mycobacterium tuberculosis* spreading to the vertebral column.

In spinal tuberculosis, the thoracic and lumbosacral segments of the vertebral column are most frequently affected. *Mycobacterium tuberculosis* dominantly affects the central and anterior regions of the vertebral bodies. Subsequently, there is considerable vertebral destruction, development of deformity, and formation of a cold abscess. Although any part of the vertebral column can be affected, the thoracic region is the most frequent. Paravertebral pus collection is often demonstrated in the thoracic vertebral region as a posterior mediastinal mass and often simulates malignancy.^3,4^ In many cases, vertebral column involvement can be a manifestation of disseminated tuberculosis/ miliary tuberculosis.^5,6^

Many isolated case reports suggest that varied thoracic imaging is common in patients with spinal tuberculosis.^7^ For example, Fennira and colleagues presented a series of 5 spinal tuberculosis cases from Tunisia. All had unusual thoracic abnormalities. In one case, the para-mediastinal abscess invaded the left lung. In other cases, thoracic imaging revealed chest wall abscess, tuberculous empyema, and miliary tuberculosis.^8^

Gupta and coworkers noted unsuspected axial skeletal tuberculosis in computed tomography studies, that were done to evaluate abdominal or pulmonary tuberculosis. The authors evaluated 726 computed tomography (CT) studies and noted unsuspected axial skeletal tuberculosis in 34 (4.7%) patients. The thoracic spine was the most commonly affected site.^9^

In this prospective study, we assessed thoracic computed tomographic abnormalities in patients with spinal tuberculosis.

## Material and methods

This was a prospective study conducted in the Department of Neurology, King George’s Medical University, Lucknow. Written consent was taken from the patient or their relatives before enrolment in the study. The study was approved by the Institutional Ethics Committee.

## Inclusion criteria

All newly diagnosed patients with spinal tuberculosis were included. Patients were diagnosed based on radiological, microbiological, and histopathological criteria. The diagnosis of spinal tuberculosis was made if the patient clinically presented with localized back pain, constitutional symptoms, localized tenderness with/without deformity, cold abscess, and/ or transverse myelopathy. The magnetic resonance imaging revealed erosion or destruction of two or more contiguous vertebral bodies and intervening discs, disc infection, along a paravertebral mass lesion or abscess. Definite spinal tuberculosis was defined when the presence of *Mycobacterium tuberculosis* was demonstrated in biopsied tissue by Xpert MTB/RIF assay or microscopy. The diagnosis was considered confirmed if histopathology demonstrated characteristic tuberculous granuloma. A tuberculous granuloma was characterized by caseation necrosis, lymphocytic or histiocytic cell infiltrations, epithelioid, and Langhan’s type giant cells. Clinical or radiological improvement following antituberculosis treatment was also considered as diagnostic criteria suggestive of tuberculous etiology.^10–12^

## Exclusion criteria

All patients with diagnosed tuberculosis of any site and already on ATT were excluded. Patients with known malignancies were excluded.

## Work up

After detailed clinical evaluation, all patients underwent hematological and biochemical tests. Testing included complete blood count, blood glucose, electrolytes, liver function test, kidney function test, and human immunodeficiency virus enzyme-linked immunosorbent assay. All patients were subjected to gadolinium-enhanced magnetic resonance imaging. MRI was done on a Signa Excite 1.5 T machine (GE Medical Systems, Milwaukee, WI, USA). They underwent CT thorax (Phillips brilliance 64 slice CT scan machine) also. CT thorax images were interpreted by an expert radiologist, who was blinded to the clinical details.

CT-guided vertebral biopsy was done and samples were collected in normal saline and formalin, in separate sterile containers. The sample preserved in formalin was sent for histopathology. A normal saline sample was sent for microbiological evaluation.

## Definitions

The thoracic computed tomographic imaging findings were categorized into parenchymal, pleural, and mediastinal categories. Lung parenchymal lesions of size <3 mm in diameter were defined as micronodules, 3-mm to 3-cm sized lesions were nodules, and lesions more than 3cm in size were defined as a lung mass. Centrilobular nodules were well-demarcated small 5-10 mm lung nodules which are anatomically located centrally, 2 mm away from the pleural surface or interlobar septa. The ‘tree in bud appearance was characterized by the presence of peripheral lesions of 2-4 mm and located within 5 mm of the pleural surface. The miliary pattern was characterized by numerable, small 1-4 mm nodular lesions seen throughout the lung field. Consolidation was defined as one or more fairly homogeneous opaque lesions leading to obscuration of the pulmonary vessels without loss of lung volume. Homogeneous opacities that do not obscure the underlying pulmonary vessels were ground-glass opacities. Lung fibrosis was diagnosed if the CT thorax demonstrated a honeycomb appearance, with or without traction bronchiectasis. Hilar lymphadenopathy was noted if the lymph node size was greater than 10 mm in diameter.^13–15^

## Treatment

As per the standard recommendation of the World Health Organisation, antituberculous treatment was given to all the patients. In the initial intensive phase (two months), the patients were given oral isoniazid (5 mg/day; maximum dose of 300 mg/day), rifampicin (10 mg/kg/day; maximum dose of 600 mg/day), pyrazinamide (25 mg/kg; maximum dose of 2000 mg/ day) and ethambutol (15 mg /kg; maximum 1200 mg). In the continuation phase, oral isoniazid and rifampicin were further continued for the next 10 months.^16^

## Follow up

The patients were followed up for the next 6 months. The outcome was assessed by using a modified Rankin scale at 6 months.

## Statistical analysis

Data were analyzed using SPSS version 24. Continuous variables were shown as mean ± standard deviation or median with range. Categorical variables were shown as a percentage. The continuous variables were compared by independent t-test. The categorical variables were compared by using the chi-square test and Fisher’s exact test. Non-parametric tests (Kruskal–Wallis test or Mann–Whitney U-test) were used for analyzing not normally distributed data. The differences were considered statistically significant if the p-value was <0.05.

## Results

In this study, we enrolled 66 patients. CT-guided biopsies were done in 24 cases. In 7 cases, histopathology demonstrated tuberculous granuloma. Only in one patient, GeneXpert MTB/RIF was positive for *Mycobacterium tuberculosis*. In the remaining 58 patients, the diagnosis of spinal tuberculosis was based on clinical and imaging findings.

On neuroimaging, the commonest vertebral column segment involved was thoracic (38 patients; 57.6%), followed by the lumbosacral segment (24 patients; 36.3%). Cervical segments were involved in 14 (21.2%)patients. In 10 patients, multiple vertebral segments were affected.

CT thorax revealed lung abnormalities in 22 patients (33.3%). Parenchymal fibrosis was the most frequent lung abnormality, that was noted in 13 patients. Six patients had lobar consolidation. In one patient each, either cavitation, atelectasis, emphysema, endobronchial tuberculosis, or traction bronchiectasis was noted. One patient had miliary mottling of the lungs. Pleural effusion was noted in 9 patients. Pleural thickening and empyema were noted in 2 patients each. One patient had hydropneumothorax. Mediastinal lymphadenopathy was noted in 3 patients. (Figure-1; Figure-2; Figure-3; Figure-4; Figure-5; Figure-6; Figure-7; Figure-8) (Table-1; Table-2)

**Figure-1.**
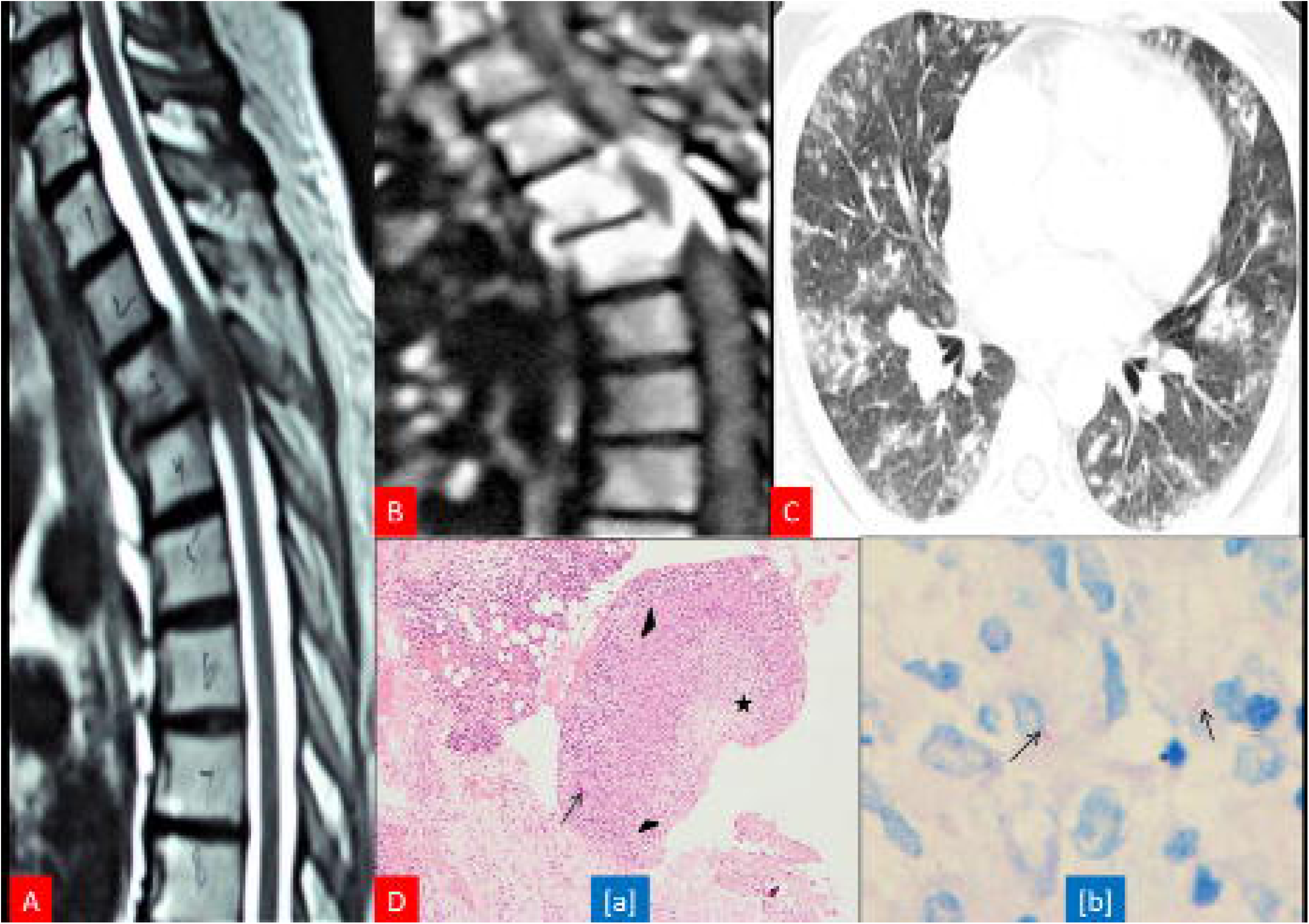
Magnetic resonance imaging of the vertebral column revealed tuberculous involvement of thoracic 3 and four segments (A and B). Computed tomography thorax shows pulmonary centrilobular nodular opacities with a tree-in-bud pattern (C). Histopathology of biopsied vertebral tissue reveals classical tuberculous granuloma. There is central caseous necrosis surrounded by epithelioid cells in Langhan’s giant cells (Hematoxylin & Eosin, x50). The section shows the presence of acid-fast bacilli on Ziehl–Neelsen staining (D).

**Figure-2.**
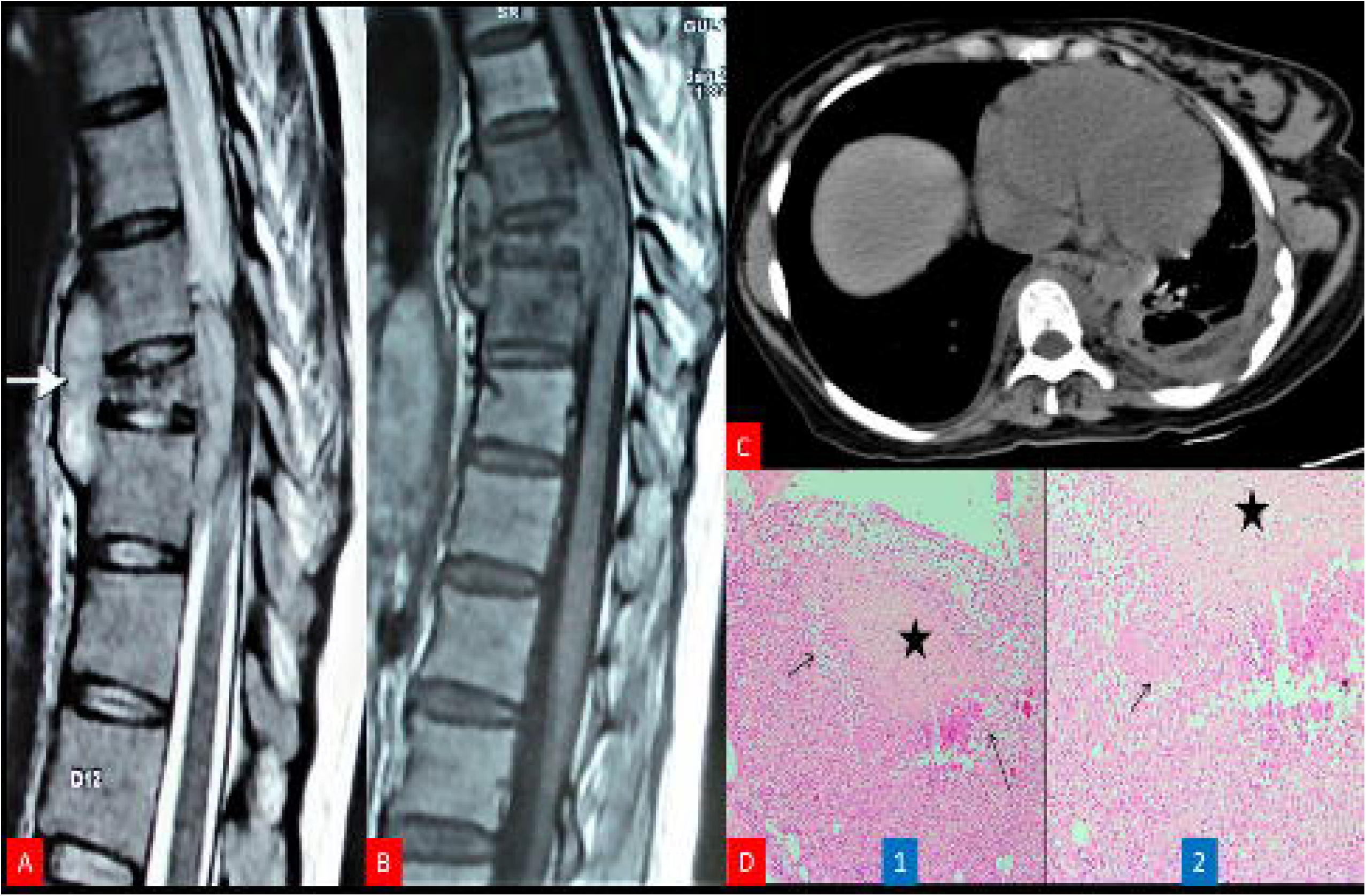
Magnetic resonance imaging of the vertebral column reveals the collapse of thoracic 9 vertebrae. Pus collection has been noted in anterior prevertebral, and anterior epidural space (A and B). Computed tomography thorax shows left-sided pleural thickening with mild pleural effusion. There is subsegmental atelectasis of underlying lung parenchyma (C). Vertebral column biopsy shows central caseous necrosis, epithelioid cells, and lymphocyte collection. (Hematoxylin & Eosin, x50) (D).

**Figure-3.**
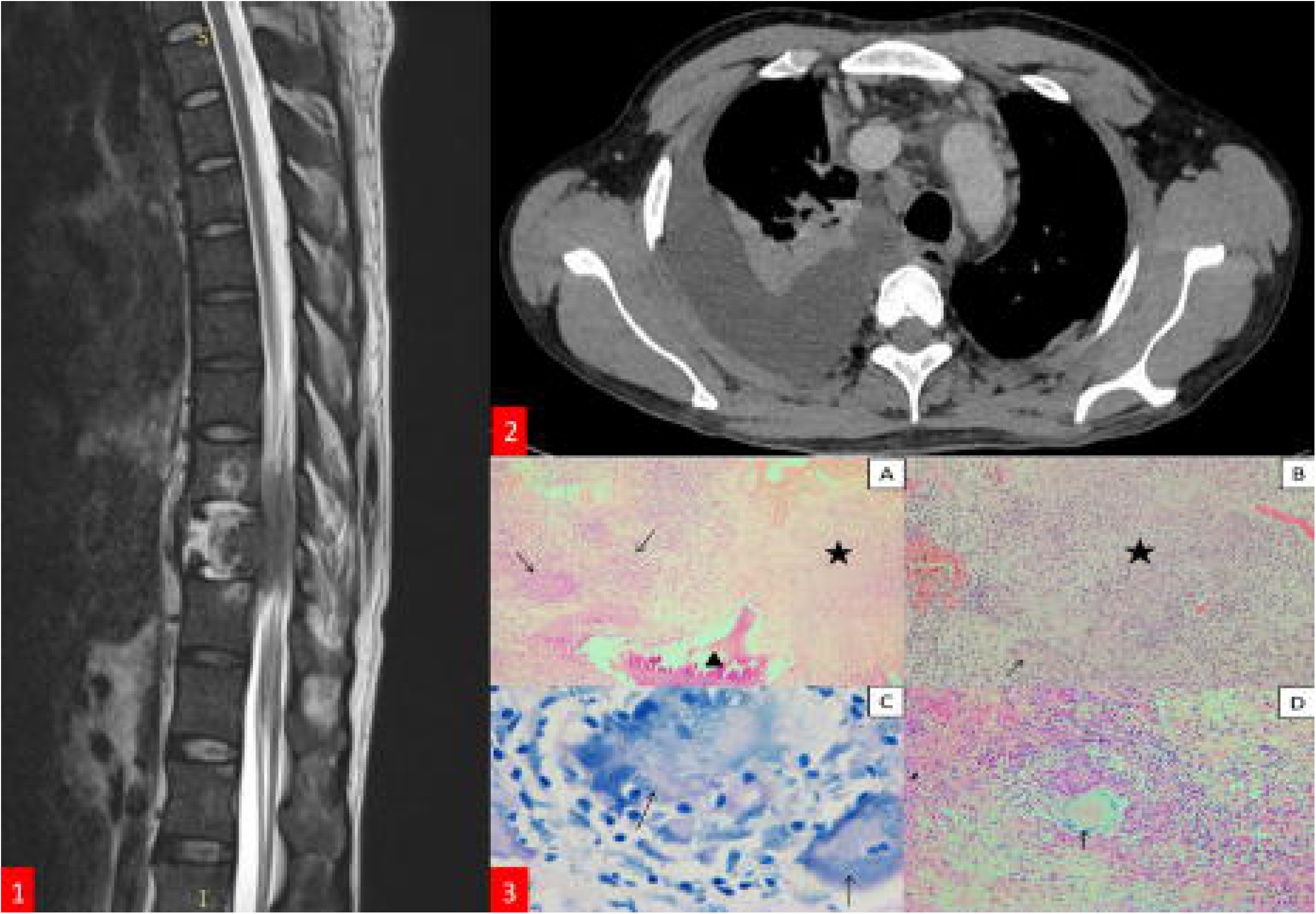
Magnetic resonance imaging of the vertebral column shows tuberculous involvement of thoracic 9-11 vertebral bodies. The paravertebral abscess was noted at the thoracic 10 levels (Figure 1). Computed tomography chest depicts right-sided pleural effusion with underlying lung collapse (Figure 2). Large conglomerate upper right paratracheal lymph nodes are noted. Histopathology of vertebral biopsied tissue shows tuberculous granuloma (Figure 3).

**Figure-4.**
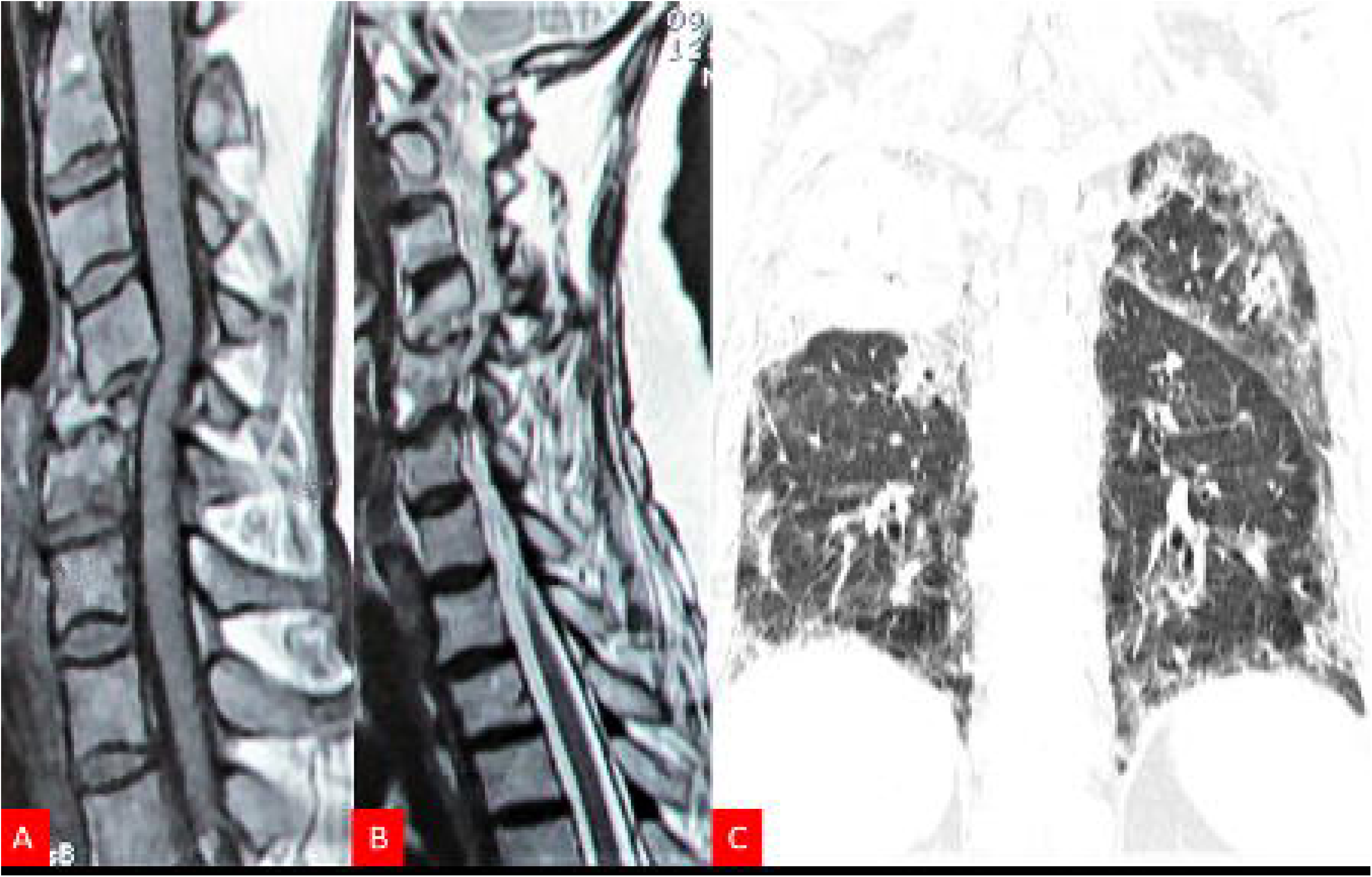
Magnetic resonance imaging of the vertebral column shows tuberculous involvement of cervical 4,5 and 6 involvement (A and B). Computed tomography chest depicts fibro-atelectasis in the right upper lobe. Multiple patchy areas of fibrosis are seen in the rest of the lung field (C).

**Figure-5.**
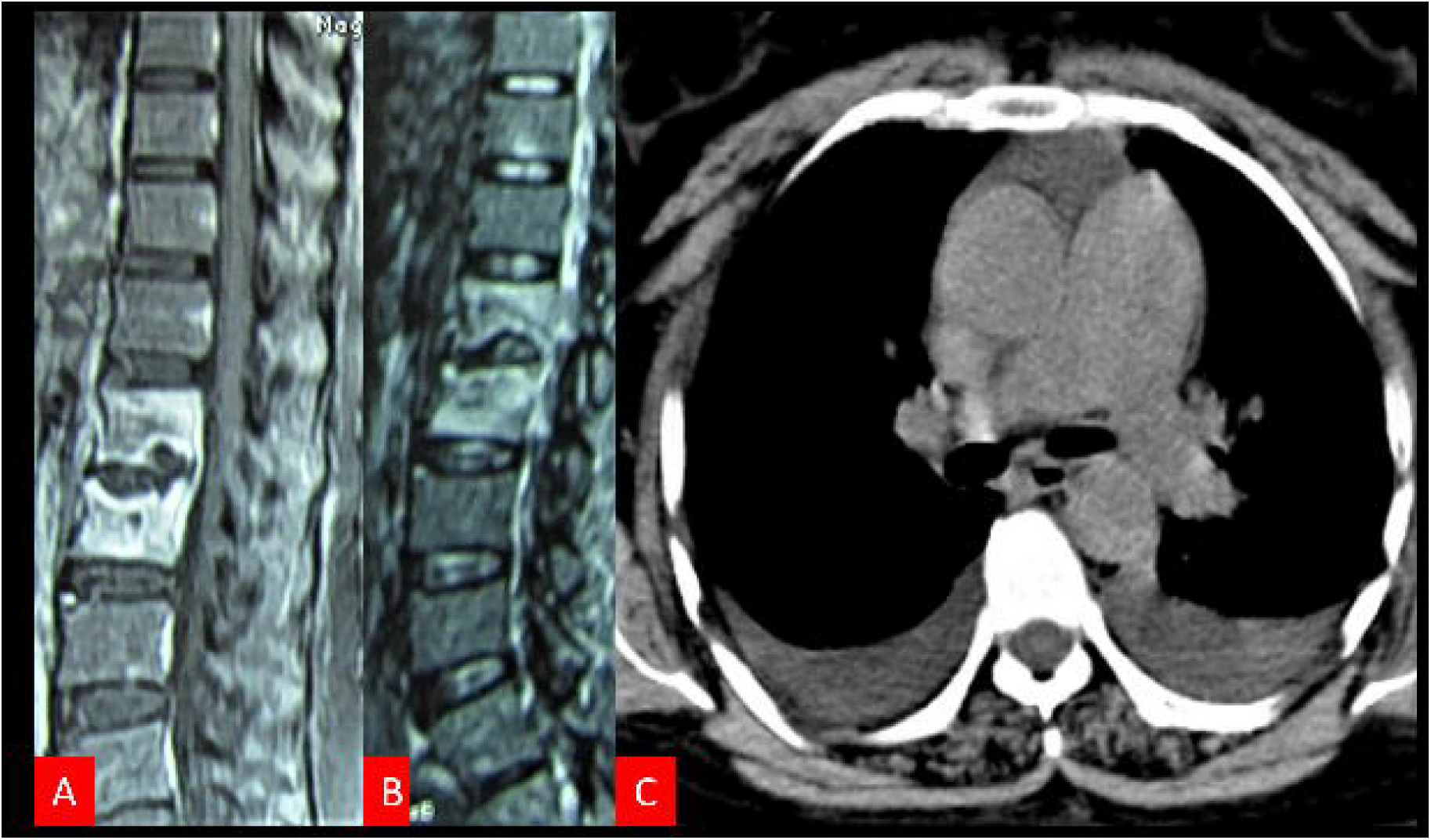
Magnetic resonance imaging the of vertebral column shows tuberculous involvement of thoracic 12 and lumbar 1 vertebral bodies (A and B). Computed tomography chest depicts bilateral pleural effusion with segmental atelectasis (C).

**Figure-6.**
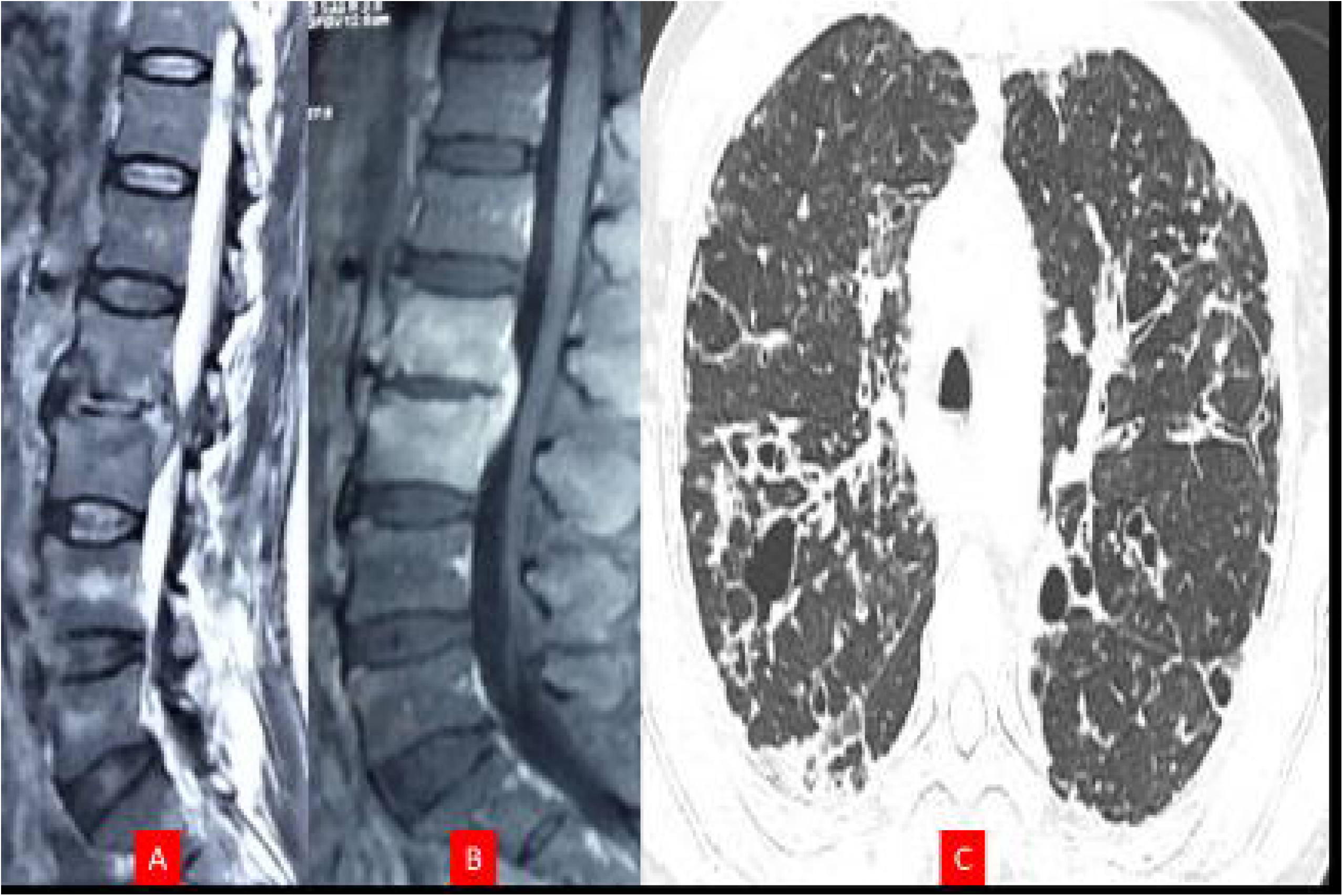
Magnetic resonance imathe ging of vertebral column shows tuberculous involvement of lumbar 2 and 3 vertebral bodies (A and B). Computed tomography thorax shows multifocal fibronodular opacities with associated traction bronchiectasis and peribronchial cuffing (C).

**Figure-7.**
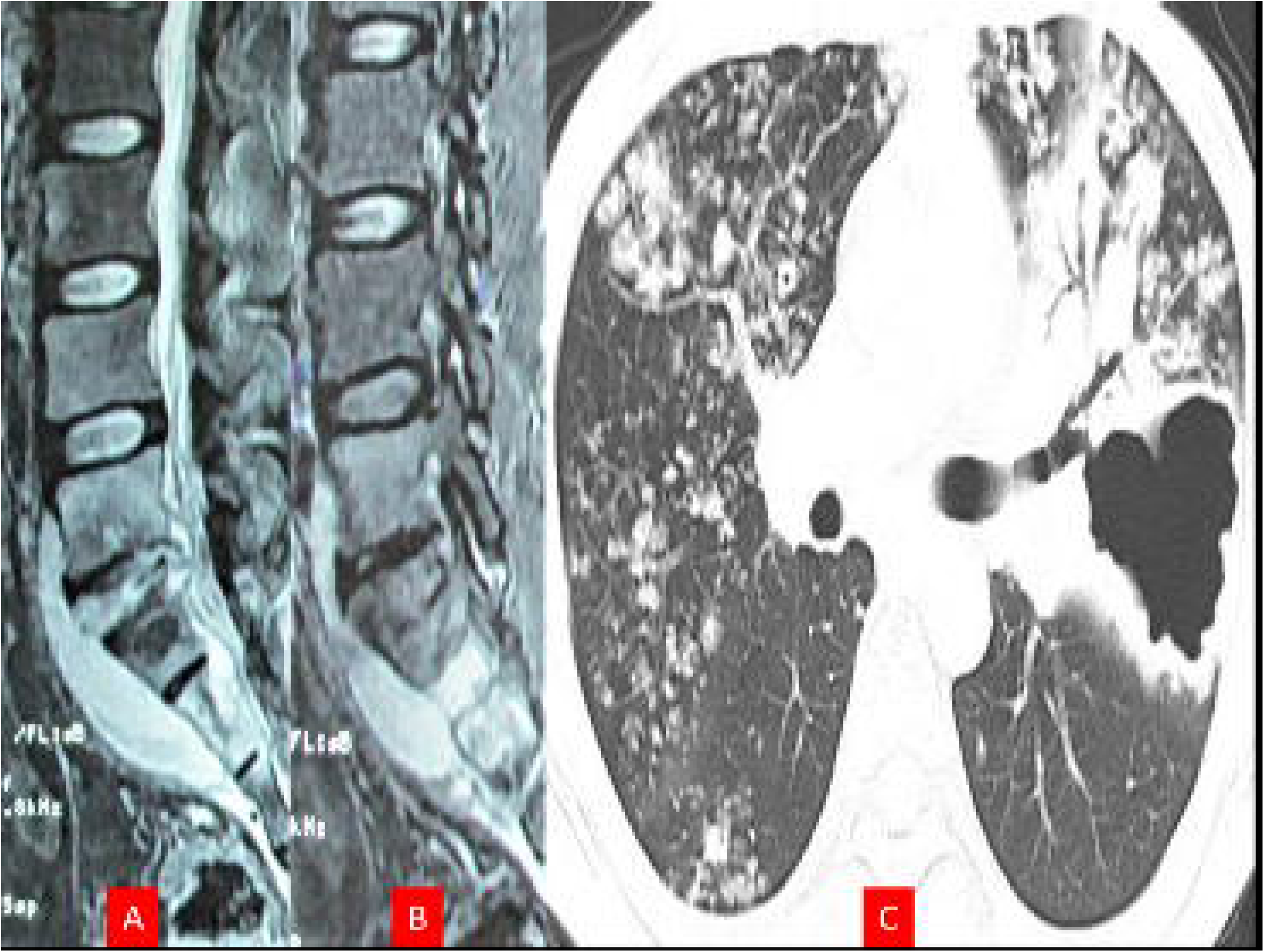
Magnetic resonance imaging of the vertebral column shows tuberculous involvement of lumbar 4 and 5 vertebral segments with para-discal involvement (A and B). Computed tomography thorax shows centrilobular and nodular opacities noted in the bilateral lung field (C).

**Figure-8.**
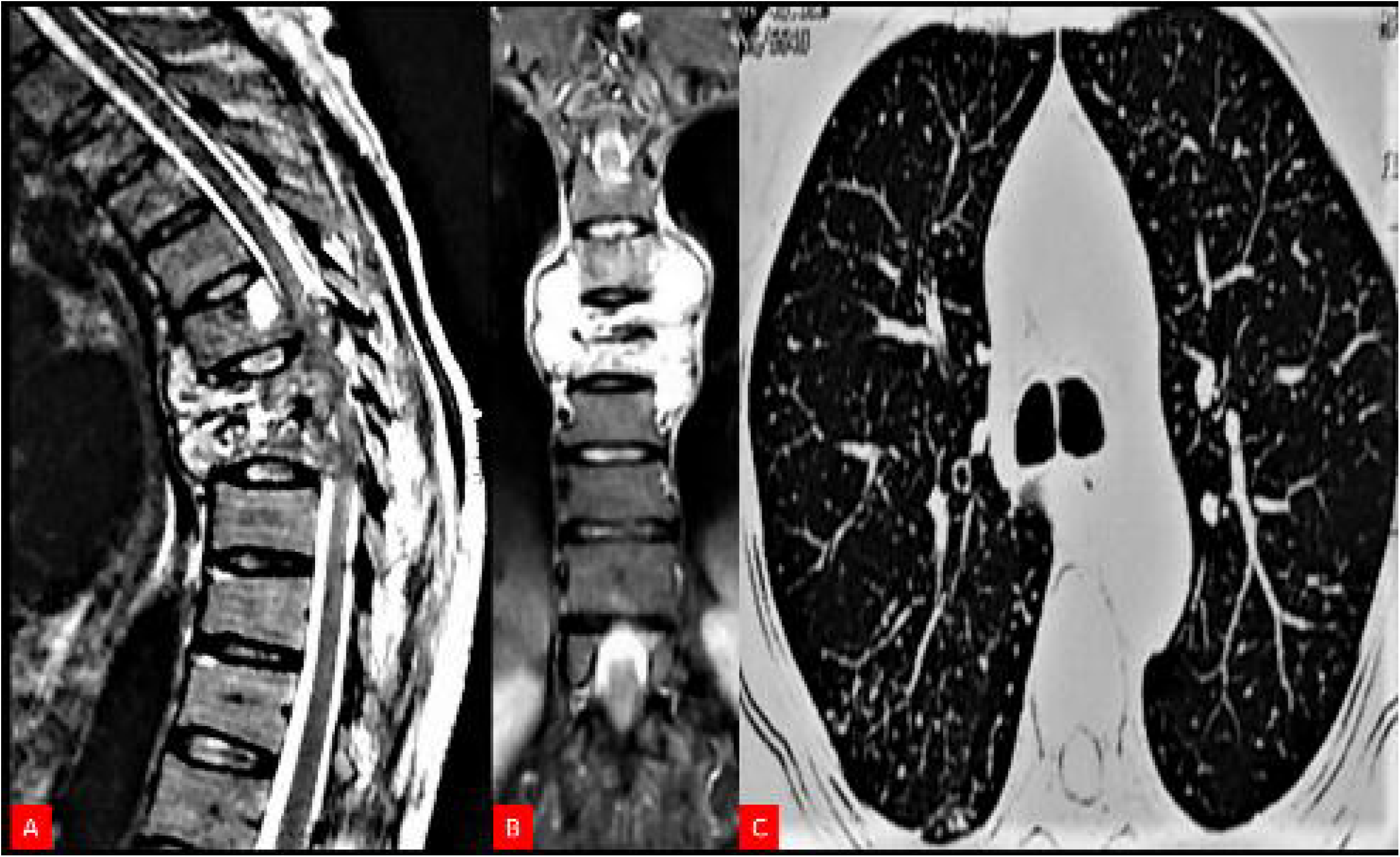
Magnetic resonance imaging of the vertebral column shows tuberculous involvement of thoracic 6,7 and 8 vertebral segments (A and B). Computed tomography thor shows miliary tuberculosis (C).

**Table-1:** Baseline characteristics of patients with spinal tuberculosis (n=66)

| Clinical variables | Total (n=66) | Patients with lung abnormalities (n=22) | Patients without lung abnormalities (n=44) | P value | Odds ratio | 95% CI |
| --- | --- | --- | --- | --- | --- | --- |
| **Gender** |  |  |  |  |  |  |
| Male | 38(57.6%) | 14(63.6%) | 24(54.5%) | 0.48 | 1.46 | [0.51-4.18] |
| Female | 28(42.4%) |  |  |  |  |  |
| **Age** |  |  |  |  |  |  |
| Mean ±SD | 38.71±15.24 | 38.73 ± 12.23 | 38.70 ±16.68 | NA | NA | NA |
| Median [IQR] | 38.00[23] | 36.5[40] | 39.00[60] |  |  |  |
| Fever | 36(54.5%) | 14(63.6%) | 22(50.0%) | 0.29 | 1.75 | [0.61-5.00] |
| Backpain | 64(97%) | 22(100%) | 42(95.5%) | 0.31 | 1.52 | [1.28-1.82] |
| Weakness of limbs | 63(95.5%) | 21(95.5%) | 42(95.5%) | 1.00 | 1.00 | [0.09-11.67] |
| Bowel and bladder involvement | 50(75.8%) | 14(63.6%) | 36(81.8%) | 0.10 | 0.39 | [0.12-1.24] |
| Sensory Symptoms | 65(98.5%) | 22(100%) | 43(97.7%) | 0.476 | 1.51 | [1.27-1.80] |
| Lymphadenopathy | 6(9.1%) | 3(13.6%) | 3(6.8%) | 0.39 | 2.16 | [0.40-11.70] |
| **Spinal magnetic resonance imaging characteristics** |  |  |  |  |  |  |
| Abnormal chest X-ray | 17(25.8%) | 12(54.5%) | 5(11.4%) | 0.001 | 9.25 | [2.83-30.29] |
| Disc involvement | 63(95.5%) | 21(95.5%) | 42(95.5%) | 1.00 | 1.00 | [0.09-11.67] |
| Epiphyseal Involvement | 58(87.9%) | 20(90.9%) | 38(86.4%) | 0.59 | 1.58 | [0.29-8.55] |
| Vertebral Body Involvement | 63(95.5%) | 22(100%) | 41(93.2%) | 0.21 | 1.54 | [0.28-1.84] |
| Pedicle involvement | 49(74.2%) | 17(77.3%) | 32(72.7%) | 0.69 | 1.28 | [0.39-4.22] |
| Paraspinal extension | 39(59.1%) | 14(63.6%) | 25(56.8%) | 0.60 | 1.33 | [0.46-3.82] |
| Adjacent soft tissue involvement | 31(47%) | 11(50.0%) | 20(45.5%) | 0.73 | 1.20 | [0.43-3.34] |

**Table-2:** Spectrum of Thoracic Computed Tomographic findings in patients with spinal tuberculosis (n=22)

| **Variables** | **Value** | **Percentage** |
| --- | --- | --- |
| **Parenchymal involvement** | 18 | 81.8% |
| Consolidation | 6 | 27.2% |
| Cavitation | 1 | 4.5% |
| Atelectasis | 1 | 4.5% |
| Miliary pattern | 1 | 4.5% |
| Fibrosis/scarring | 13 | 51.5% |
| Emphysema | 1 | 4.5% |
| Traction bronchiectasis | 1 | 4.5% |
| Endobronchial tuberculosis | 1 | 4.5% |
| **Pleural involvement** | 11 | 50% |
| Pleural effusion | 9 | 40.9% |
| Pleural thickening | 2 | 9.1% |
| Empyema | 2 | 9.1% |
| Pneumothorax | 0 | 0% |
| Hydropneumothorax | 1 | 4.5% |
| **Lymphadenopathy** | 6 | 27.3% |
| Hilar and Mediastinal lymphadenopathy | 6 | 27.3% |

## Follow up

At inclusion, 59 (89.4%) patients had modified Rankin scale scores≥3. After 6 months, 58 (87.9%) patients showed improvement and achieved a modified Rankin scale score of <3. In 4 patients, spinal surgery was needed. 3 patients did not have an improvement in disability.

Univariate analysis did not reveal significant clinical or imaging variables, that were associated with improvement in the modified Rankin scale score.

## Discussion

In our study, we noted that approximately one-third of patients with spinal tuberculosis have concomitant pathologies in the lungs as well. Lung parenchyma, pleura, and lymph nodes could all be affected. Pulmonary fibrosis and pleural effusion were the commonest lung abnormalities.

Spinal tuberculosis is a great masquerader of various lung pathologies, particularly malignancy, and frequently poses diagnostic challenges.^17^ For example, spinal tuberculosis of the thoracic region can appear as mediastinal widening. Various other causes manifesting as mediastinal widening include mediastinal lymphadenopathy, pericardial effusion, paravertebral abscess, and aneurysm of the aorta.^18–20^

Frequent lung involvement in spinal tuberculosis could be indicative of disseminated tuberculosis. Disseminated tuberculosis is characterized by the tuberculous involvement of two or more non-contiguous body organs. The presence of *Mycobacterium tuberculosis* should be there in one of the body structures. Miliary tuberculosis was characterized by the demonstration of innumerable small lung tubercles of 1–2 mm in diameter. Miliary lesions, characteristically, affect both the lungs. ^5,21^ Ringshausen et al described a case of spinal tuberculosis; this patient had a solitary pulmonary nodule. The nodule was suspected of metastatic lung cancer and was irradiated. Subsequently, the disease progressed to evolve into florid central nervous system tuberculosis.^4^ Rarely, a spinal tuberculosis-associated paravertebral abscess can mimic an aortic aneurysm. ^22^

Shim and colleagues reviewed data from 50 patients with spinal tuberculosis. These patients were additionally subjected to x-ray chest and/or thoracic computed tomography. In this series es, the authors noted that in 21 (42%) cases there were thoracic imaging abnormalities consistent with pulmonary tuberculosis were noted. The most frequent abnormality was centrilobular nodules were seen in 10 patients. Other abnormalities seen were tree-in-bud appearance, consolidation, cavitation, miliary tuberculosis, and pleural effusion or parenchymal involvement. All these forms of tuberculosis represent active disease and help in establishing tuberculous etiology. Paravertebral tuberculous abscess mostly tracks down to the mediastinum and pleural cavity. In spinal tuberculosis, lung involvement could be asymptomatic. Lung involvement in spinal tuberculosis is most frequently noted if tuberculosis is localized to the thoracic region. ^23^

The addition of thoracic CT in the evaluation of spinal tuberculosis may lead to added evidence of tuberculosis and the detection of alternate aetiologies.

Histopathological confirmation requires an adequate sample. A CT-guided biopsy is better and yields good results. In a recent study by Yu et al, GeneXpert MTB/RIF plays a complementary role with conventional histopathology.^24^

The major limitation of our study was the inability to secure microbiological confirmation in the majority of cases. The diagnosis was based on clinical characteristics, imaging findings, and response to antituberculous treatment.

In conclusion, one-third of spinal tuberculosis patients have concomitant tuberculous lung abnormalities. A lung lesion consistent with tuberculosis can help in making a more reliable diagnosis of spinal tuberculosis, in the absence of microbiological confirmation.

## Declaration of Generative AI and AI-assisted technologies in the writing process

None

## Data Availability

All data produced in the present work are contained in the manuscript.

